# Impact of Non-Standardized Reporting on Reproducibility, Usability, and Integration in Nasopharyngeal Metagenomic Research: A Systematic Review

**DOI:** 10.1101/2025.09.06.25335230

**Authors:** Monica L Bustos, Kuncheng Song, Hayden N Brochu, Qimin Zhang, Lakshmanan K Iyer, Crystal R Icenhour

## Abstract

The nasopharyngeal microbiome plays an essential role in respiratory health and disease, making it a key focus of metagenomic research. However, inconsistent reporting standards across studies hinder reproducibility, usability, and integration of these data, limiting the scientific value of nasopharyngeal metagenomic datasets. This systematic review assessed the impact of non-standardized reporting on metagenomic studies, focusing on reproducibility, usability, and integration in publicly available datasets. We screened 988 studies pertaining to research on the nasopharyngeal microbiome. Of the screened manuscripts, 227 were selected for full-text review based on detailed inclusion and exclusion criteria. Key findings included that only 78 studies (34%) had reproducible methods sections, 33 of those 78 studies (15%) provided analytically sufficient metadata, and 4% demonstrated mismatched laboratory methods incompatible with reported datasets. We attributed these inconsistencies to gaps in methodological transparency, lack of accessible metadata, and misaligned file formats, which collectively impede dataset reuse and integration. The interchangeability of nasopharyngeal aspirates (NPA) and nasopharyngeal swabs (NPS) was evaluated using reproducible datasets. Significant variation in the microbial profile between source types was confirmed, highlighting that specimen interchange would be inappropriate within a study. Our results underscore the critical need for standardized reporting guidelines in metagenomic research to improve data transparency, facilitate reproducibility, and enable broader data integration. The adoption of comprehensive and consistent reporting practices would significantly enhance the scientific utility of nasopharyngeal microbiome studies, promoting reliable, replicable, and integrative research across the field.

## Introduction

The nasopharyngeal microbiome, a diverse population of microorganisms residing in the upper respiratory tract, plays a crucial role in maintaining respiratory health and influencing disease susceptibility (1–3). As sequencing technologies advance and metagenomic approaches evolve, the nasopharyngeal microbiome has become a key focus of research, particularly in the context of infectious diseases, chronic conditions like asthma and sinusitis, and the impact of the microbiome on immune system modulation (4,5). Despite the generation of vast nasopharyngeal microbiome datasets, challenges related to data reproducibility, usability, and integration persist, impeding expansion of metagenomic research (6,7). These issues are exacerbated by the lack of standardized reporting practices across studies, which hinders the validation of findings, the reuse of data in future research, and the integration of datasets from diverse sources or studies (8). The lack of reproducible data in published metagenomic studies has significant and wide-ranging implications, including delayed or ineffective medical and public health applications, ineffective interventions, and wasted resources through duplicated research efforts (8,9).

Non-standardized reporting of metagenomic data manifests in multiple ways within nasopharyngeal microbiome studies, including inconsistent descriptions of sampling methods, variations in sequencing techniques, and inadequate metadata, such as demographic or clinical information (10). These inconsistencies complicate efforts to reproduce results, making it difficult for independent researchers to verify findings or conduct similar studies using the same methodologies (11). Furthermore, without transparent reporting of methods and data processing workflows, researchers struggle to understand how datasets were generated, hindering effective data reuse and cross-study comparisons (9). Integration of nasopharyngeal microbiome datasets from different studies is similarly hindered by these reporting challenges, limiting the capacity to draw broader conclusions about microbial diversity and its clinical relevance across populations and disease states (8).

Characterizing the healthy nasopharyngeal microbiome remains an emerging and complex undertaking, requiring a broad and diverse dataset to ensure analytical robustness and reproducibility. To support the standardization of a nasopharyngeal bioinformatic resource for genomic surveillance of respiratory pathogen outbreaks, this systematic review evaluated publicly available nasopharyngeal microbiome publications. Despite the abundance of published studies, only 34% provided reproducible data suitable for inclusion due to inconsistent reporting practices. This review identified three major barriers to effective data utilization: (1) lack of reproducibility, defined as the inability of independent researchers to replicate findings from existing datasets; (2) limited usability, or the practical challenges of accessing and applying publicly available data; and (3) poor integration potential, reflecting difficulties in synthesizing datasets across studies for comparative or systematic review purposes. Additionally, this review assessed the interchangeability of nasopharyngeal aspirates (NPA) and nasopharyngeal swabs (NPS), a common variation in study protocols. Given the distinct methodologies used for each sampling technique, the data were analyzed to determine whether both sample types yielded comparable microbiome profiles. This report aims to highlight critical gaps in standardization, encourage methodological transparency, and provide recommendations to enhance reproducibility and data integration in future nasopharyngeal microbiome research.

## Methods

### Data Sources and Search Strategy

We conducted a systematic search of the PubMed (https://pubmed.ncbi.nlm.nih.gov/) and Embase (https://www.embase.com/) (⍰databases with the primary goal of curating publicly accessible nasopharyngeal microbiome publications. This review also assessed the reproducibility and replicability of healthy nasopharyngeal whole-genome datasets, emphasizing the impact of non-standardized reporting on their usability and integration in future research. The search strategy employed (**Figure 1**), with the search restricted to studies published up to May 18, 2024. The initial query returned 988 records, which were first deduplicated to remove overlapping studies between databases. The remaining unique records were then subjected to a title and abstract screening by two independent reviewers (MB and KS). This screening process applied strict eligibility criteria to ensure manuscripts represented novel research studies, employed appropriate next-generation sequencing techniques, utilized nasopharyngeal sampling methods, analyzed human subjects, were available in English, and used Illumina sequencing platforms. Studies that did not meet these criteria were excluded. When eligibility was unclear or reviewers disagreed, a third reviewer (CI) resolved the discrepancy. Publications passing initial screening underwent full-text review to confirm adherence to all inclusion and exclusion criteria. The screening process is summarized in **Figure 1**. Of the 227 studies that met the initial inclusion criteria, 78 were ultimately selected for dataset download and further reprocessing through our standardized bioinformatics pipeline. This meta-analysis curation process was also included in our independent study by Song et al.(1), which provides a deeper analysis of the nasopharyngeal microbiome to detail the creation of an analytical framework that allows for the translation of nasopharyngeal microbiome data into clinically actionable results. These studies were selected based on the availability and quality of accompanying metadata, which was then evaluated using several key criteria: clear differentiation between control and disease states, accurate alignment of reported methods with raw data files, metadata completeness, and the identification of baseline data in longitudinal designs. These requirements ensured that selected datasets were structured to support robust and reproducible downstream analysis.

**Figure 1.**
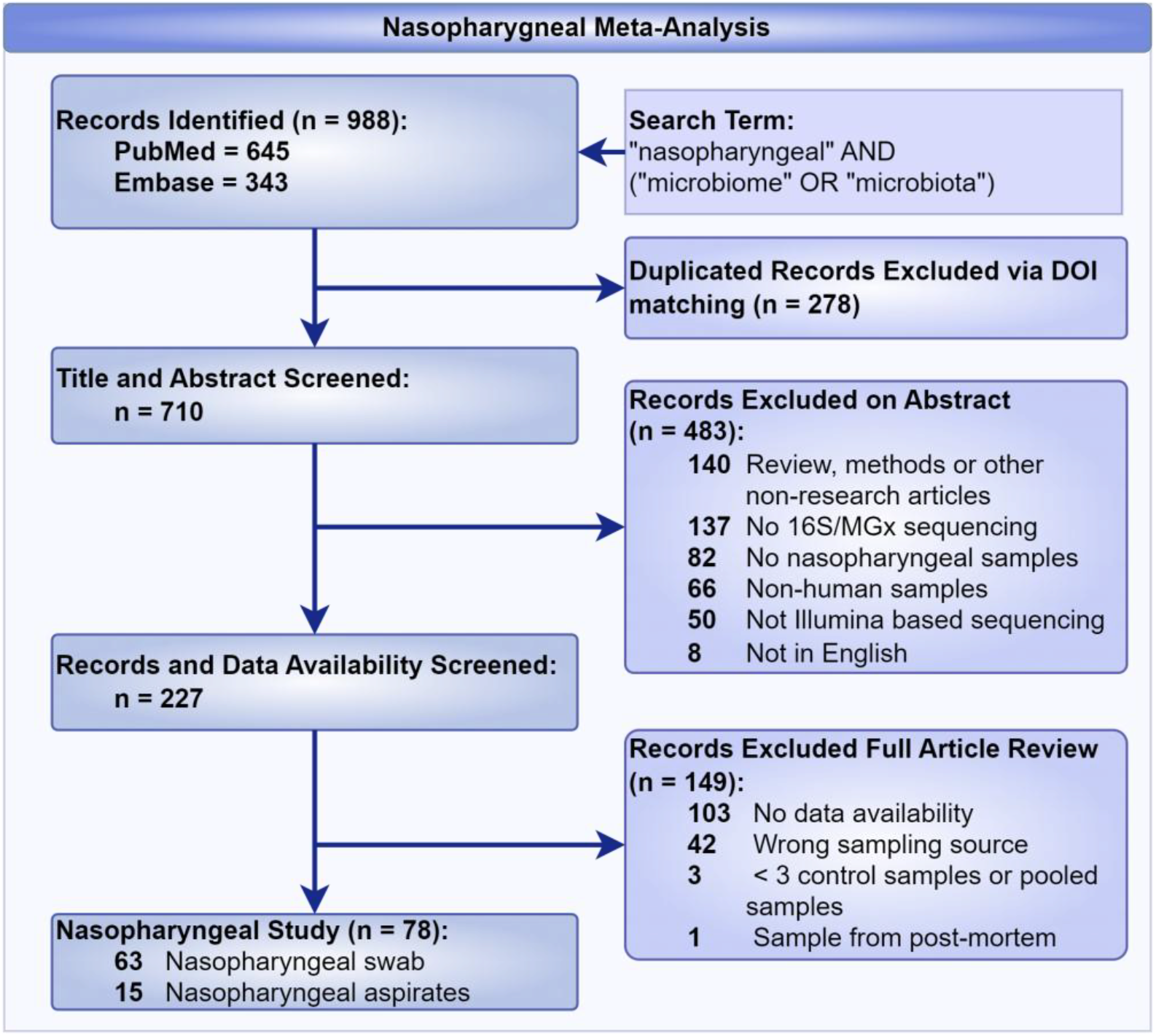
Preferred Reporting Items for Systematic Review for the Curation of a Healthy Nasopharyngeal Bioinformatic Library

During the meta-analysis conducted for the curation of publicly available healthy nasopharyngeal microbiome data, a significant portion of the initial records were excluded due to issues with data accessibility and methodological reporting. Out of 227 manuscripts selected for full article review, 149 were excluded, primarily because of inaccessible data or discrepancies within the methods section. These limitations in data availability and reporting highlighted critical challenges in ensuring reproducibility and reliability, which directly influenced the inclusion and exclusion criteria for this review. This finding led the reviewers to recognize potential broader issues with methodological reproducibility, prompting a secondary review focused on assessing the replicability of the methodologies employed. Of the 227 records, 78 articles were identified as having nasopharyngeal sources, which were subsequently selected for a more rigorous evaluation of their methodology. The criteria used in this process are outlined below.

### Inclusion and Exclusion Criteria

Inclusion Criteria (studies were included in the meta-analysis if they met the following criteria):

1. Provided metagenomic sequencing data in a publicly accessible format.
2. Focused on nasopharyngeal specimen collection.
3. Reported relevant outcomes, such as microbial diversity or specific taxonomic abundances.

Exclusion Criteria (studies were excluded based on the following criteria):

1. Lack of data availability (including original datasets, publicly available datasets, and Illumina-based 16S/metagenomic studies).
2. Absence of clearly defined nasopharyngeal specimen collection.
3. Fewer than three unique subjects (data could not be pooled across sources or individuals).
4. Inclusion of post-mortem samples.

### Quality Assessment

The quality of the included studies was evaluated based on study design, sample size, data completeness, and reporting quality. Selected dataset quality assessment is documented in the next section.

### DADA2 Processing, Quality Control, and Taxonomic Analysis

Raw 16S V4 rRNA gene sequencing data were obtained from NCBI BioProject PRJNA997934 (nasopharyngeal swabs, NPS) and 16S V3V4 rRNA gene sequencing data from PRJNA275918 (nasopharyngeal aspirates, NPA) using fasterq-dump from the NCBI SRA Toolkit v3.1.0 (12). To match the infant-only cohort in the NPA dataset (age <1 year), NPS samples were filtered accordingly. Only samples labeled as healthy were retained from both datasets.

Read processing was performed in R v4.1.1 (13). For the NPA dataset (V3V4 region), reads were reoriented when necessary. All reads were then trimmed using the filterAndTrim function in DADA2 v1.22.0 (14). Read lengths were 2×250 bp (NPS) and 2×150 bp (NPA), with primers excluded. An in-house R function optimized trimming based on quality profiles, primer presence, and expected insert length (~256 bp). Trim parameters were set to trimLeft = c(10,10) and trimRight = c(40,189) for NPS, and c(20,19) for NPA.

Quality-filtered reads underwent standard DADA2 processing for denoising, merging, and generation of amplicon sequence variants (ASVs). Singletons and ASVs <350 bp were removed. Taxonomic classification was performed using the SILVA v138.2 database (15) with the DADA2 assignTaxonomy (minBoot = 80) and addSpecies functions. ASVs lacking at least family-level classification were excluded. Remaining ASVs were aggregated to their lowest assigned taxonomic rank to generate the count matrix.

To mitigate batch effects, taxa commonly associated with laboratory or environmental contamination (e.g., kit/reagents origin microbes known collectively as “kitome,” marine, or soil microbes) were removed. Only samples with ≥5,000 reads were retained, yielding the final aggregated count matrix for downstream analysis.

### Alpha Diversity, Dissimilarity, and Differential Abundance Analyses

Alpha diversity was calculated using the Shannon index via the diversity function (vegan v2.6-2) (16). Rarefaction was performed across thresholds from 5,000 to 100,000 reads (in 5,000-read increments) using the rrarefy function, repeated five times per threshold. Median values were used per sample, and Wilcoxon rank-sum tests assessed diversity differences between groups. Community composition differences were evaluated using Bray-Curtis dissimilarity and visualized via multidimensional scaling (MDS) with cmdscale. Statistical significance of clustering was tested using PERMANOVA (adonis2) with 999 permutations. Differential abundance (DA) analysis between NPS and NPA groups was conducted in R v4.4.3 (13) using ancombc2 (ANCOM-BC v2.8.1) (17) with input formatted via phyloseq v1.50.0 (18). Taxa with FDR-adjusted *P* < 0.05 were considered significant. Sensitivity to pseudo-count selection was noted. Taxa with positive or negative log2 fold-change (L2FC) were classified as enriched or depleted, respectively.

### Statistical Analysis

False discovery rate (FDR) correction was applied via the Benjamini-Hochberg method for all multiple hypothesis testing. Co-exclusive microbial detections were evaluated using Pearson’s Chi-squared test.

### Data Availability

All datasets included in this meta-analysis are publicly available from NCBI SRA, and links to the datasets can be found in the supplementary materials section.

## Results

In this systematic review, we initially screened 988 manuscripts by abstract and title, ultimately selecting 227 for full-text review. Upon further examination, 148 manuscripts contained appropriate qualifying nasopharyngeal sources for curating publicly available healthy nasopharyngeal bioinformatics data. However, only 78 articles contained reproducible methods sections, as assessed using the screening criteria outlined in **Figure 2**. The secondary review evaluated the impact of non-standardized reporting on the reproducibility, usability, and integration of metagenomic datasets in subsequent research. The studies were selected based on criteria that mandated the use of metagenomic sequencing to characterize the nasopharyngeal microbiome, with a specific focus on challenges related to reproducibility, replicability, clarity of terminology, usability, and the integration of datasets across studies. The results are structured around three principal themes: reproducibility, usability, and the integration of nasopharyngeal microbiome datasets.

**Figure 2.**
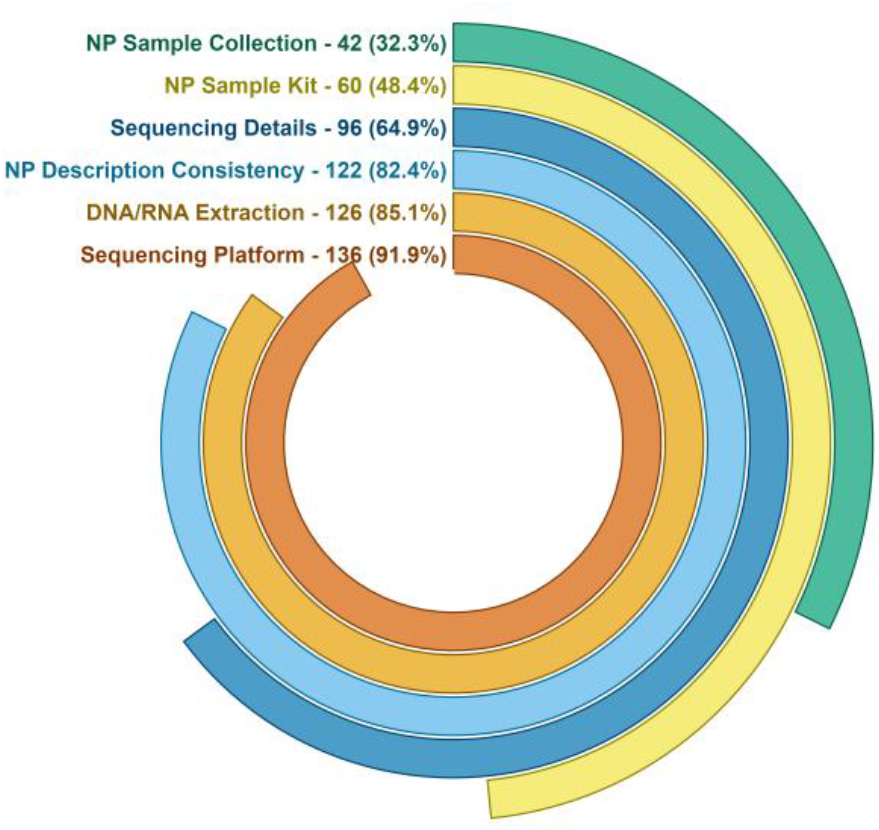
Percentage of publications with reproducible methodologies. The number of studies and percentage across different reproducibility were displayed for the 148 screened publications.

### Reproducibility

Our evaluation of methodology across the 148 selected studies revealed significant reporting gaps that hindered reproducibility and comparability of nasopharyngeal microbiome research. These gaps were particularly evident in three key areas: sampling methods, sequencing and extraction methods, and overall methodological transparency. Many studies lacked sufficient detail regarding nasopharyngeal sampling procedures, with variations in collection protocols and storage conditions, making it difficult to replicate studies or compare results across cohorts.

To assess data acceptability, we reviewed the methodology of 148 studies (**Figure 2**). A significant proportion lacked sufficient detail in their nasopharyngeal sampling procedures (n=102, 68.9%) or exhibited inconsistencies in sample collection protocols (n=26, 17.6%). These inconsistencies included variations in the method of collection, storage conditions, and the specific sources from which samples were collected. Furthermore, 88 studies (59.5%) did not report essential information regarding the collection kits used, such as the type of swab collection kit or the transport medium, leaving critical gaps in the methodology. These shortcomings posed substantial challenges for replicating studies or comparing results across cohorts. For instance, variations in the type of swab used (e.g., nasal swabs versus nasopharyngeal swabs) and discrepancies in sample preservation methods led to differences in microbial profiles, making it difficult to reproduce findings and draw consistent conclusions. These issues underscore the need for more standardized and transparent reporting in microbiome research to ensure the reliability and comparability of results across studies.

Additionally, many studies failed to specify critical details about extraction kits, sequencing procedures, and associated variables such as read depth, contributing to discrepancies in microbiome characterization. Among the 148 studies reviewed for methodological transparency, 22 studies (14.9%) did not specify the extraction method or kit for isolating DNA/RNA from nasopharyngeal samples. This lack of information is concerning, as variations in extraction protocols can significantly impact the quality and composition of the extracted DNA/RNA, thereby influencing subsequent analyses. In addition, 52 studies (35.1%) failed to disclose key sequencing procedures, such as the sequencing format used during 16S sequencing (i.e., PCR primers and hypervariable region(s) targeted). Sequencing format is a critical parameter that can affect the accuracy and resolution of taxonomic and functional precision of the detected microbes. Without this information, assessing the reliability of reported microbial profiles and performing cross-study comparisons becomes difficult. These omissions highlight the importance of detailed and standardized reporting in sequencing methodologies, as even minor differences in extraction and sequencing techniques can lead to substantial discrepancies in microbiome data.

Finally, many studies lacked transparency regarding control samples, sequencing platform-specific biases, and software versions, all of which are essential for ensuring the reproducibility of findings. Twelve studies (8.1%) failed to report critical methodological details essential for reproducibility and comparability of results. These studies omitted information about control samples, sequencing platform-specific biases, and/or software versions used in their analyses. Such omissions are particularly concerning given the diversity of sequencing platforms currently available, each with its own set of potential biases that can significantly influence microbiome study outcomes. Furthermore, the use of different taxonomic databases across studies added another layer of complexity. Variations in database selection, combined with inconsistent thresholds for taxonomic classification, contributed substantially to discrepancies in reported microbiome compositions. These inconsistencies create barriers to direct comparison between studies and complicate result interpretation. The lack of transparency regarding these methodological choices highlights the urgent need for standardization in nasopharyngeal microbiome research to facilitate reliable cross-study comparisons.

Of the 148 studies selected for methodology review, we identified 78 with sufficiently reproducible methods sections, which we subsequently selected for dataset download and metadata analysis. Despite this careful selection, the lack of standardized sampling methods, sequencing protocols, and bioinformatics workflows significantly impeded reproducibility across nasopharyngeal microbiome studies. Inconsistencies in methodological reporting directly contributed to variations in microbial composition across studies, making independent replication difficult or impossible in many cases. These methodological discrepancies underscore the critical need for greater standardization in both the design and reporting of microbiome studies to ensure reliable, reproducible findings that can advance the field.

### Usability

Of the 148 manuscripts reviewed for methodology, we identified only 78 with sufficiently reproducible methods sections for dataset download and metadata analysis. While all 78 studies provided publicly available data, significant usability issues hindered their practical application in further research. As **Figure 3** illustrates, 45 studies (57.7%) exhibited usability challenges, primarily related to difficulties in accessing, interpreting, and utilizing the metadata associated with the original datasets. A frequent issue was the inadequate reporting of participant characteristics, such as age, gender, disease status, and antibiotic usage, all essential for contextualizing microbiome data in clinical or ecological studies. Specifically, two studies (3.1%) did not identify sources in multi-source studies, while nine studies (14.1%) listed sources that conflicted with their methods sections. Furthermore, 28 studies (43.8%) omitted disease status information, precluding differentiation between control and experimental groups, and two studies (3.1%) lacked baseline identifiers for longitudinal analyses. The absence of these critical clinical and demographic variables limited the ability to assess the relevance of the microbiome findings or to determine whether the results could be generalized to other populations or contexts. Consequently, despite data availability, these usability issues substantially reduced the value of the datasets for secondary analyses and replication efforts. Insufficient metadata and difficulties in data access significantly limited the usability of nasopharyngeal microbiome datasets. The lack of transparency in data sharing further restricted the potential for secondary analysis and wider dissemination of findings.

**Figure 3.**
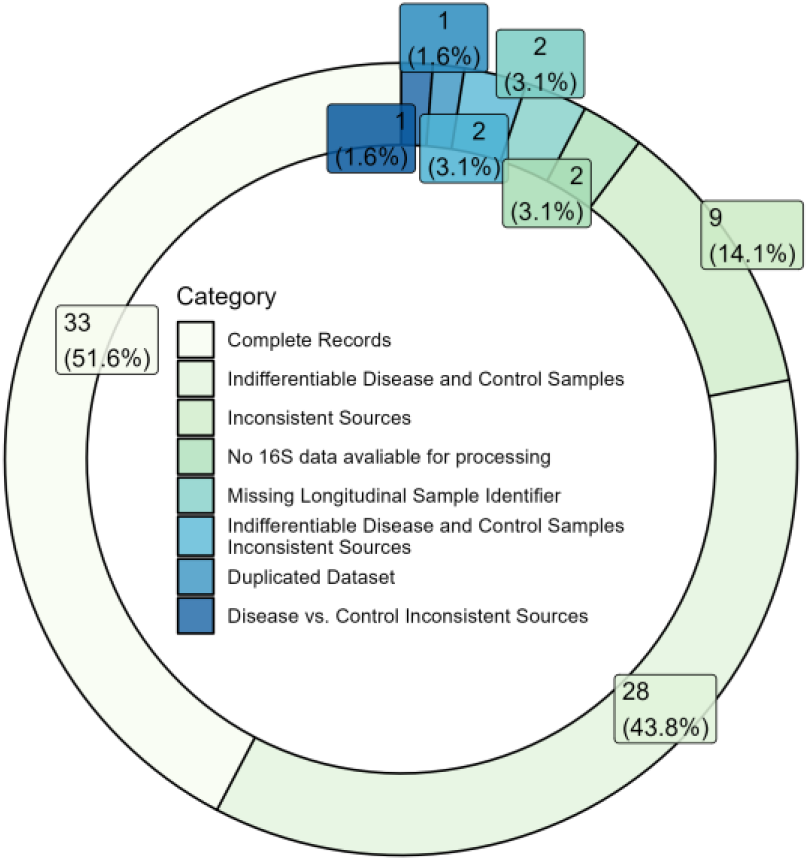
Percentage of publications with discrepancies in metadata. The number of studies and percentages are displayed for each category.

### Integration

We identified integration issues in 2 studies (3.1%), primarily stemming from misidentification of sequencing file types. These studies reported using one sequencing platform in their methods sections (e.g., Illumina) but instead employed a different platform (e.g., 454), resulting in unexpected file formats. This discrepancy creates significant technical barriers because bioinformatic pipelines are typically designed for specific file types (e.g., FASTQ or BAM) and cannot process misidentified file types without substantial modification. Aside from technical challenges, these inconsistencies between reported and actual sequencing platforms compromise data integrity and reproducibility. Such integration barriers effectively prevent large-scale meta-analyses and comparative studies that could otherwise provide comprehensive insights into the role of the nasopharyngeal microbiome in health and disease.

### Sampling Discrepancies for the Nasopharyngeal Source

Although commonly occurring in nasopharyngeal studies, the alternating method of nasopharyngeal sampling, whether through aspirate (NPA) or swab (NPS), may influence the microbial profiles detected by sequencing. To explore this, we analyzed publicly available NPA and NPS datasets, focusing on healthy infants to control for age and health status, and compared the microbial diversity and composition between the two methods..

Raw 16S rRNA gene sequencing data were processed using DADA2 to generate amplicon sequence variants (ASVs) aggregated taxonomically to reduce sparsity. Given that the studies were conducted in different laboratories using different protocols, we applied stringent background filtering to remove potential contaminants. Of 1,240 initial taxa, 608 (~49%) were identified as background. Despite their number, these taxa were rare, with median relative abundances (RAs) of 0.3% (NPA) and 0.7% (NPS) (**Figure 4a**). Their removal reduced batch effects by eliminating taxa with disproportionately high prevalence in one dataset, such as *f_Bacillaceae* (95% NPA vs. 7% NPS) and *g_Ralstonia* (0.3% NPA vs. 58% NPS) (**Figure 4b**). After filtering and retaining only samples with ≥5,000 reads, we obtained a final dataset of 632 taxa across 317 NPS and 391 NPA samples.

**Figure 4.**
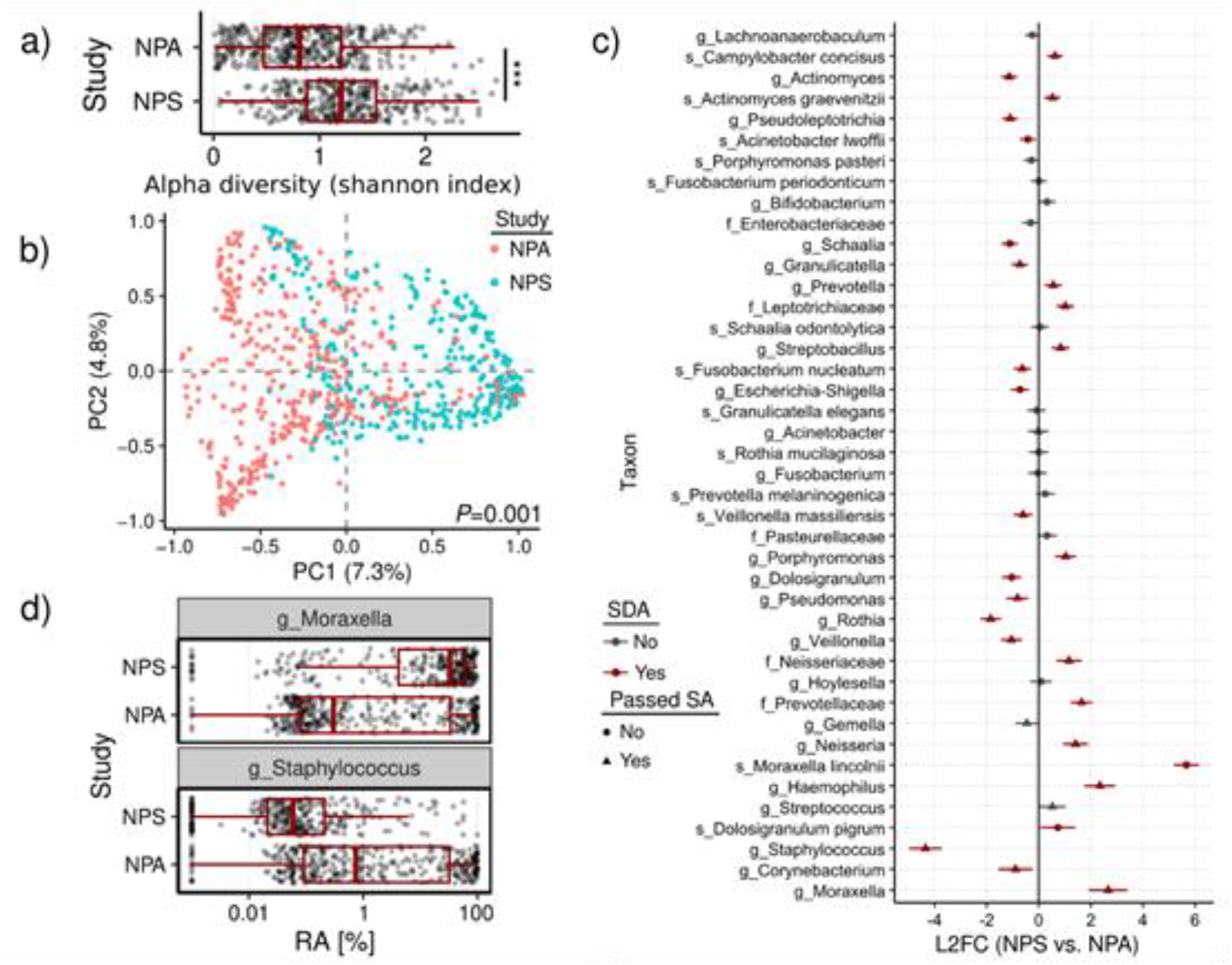
Comparison of nasopharyngeal swab (NPS) and aspirate (NPA) microbiomes. **a)** Alpha diversity computed using the Shannon index compared across studies with boxplots overlayed by jittered sample data points. **b)** Multidimensional scaling (MDS) of pairwise Bray-Curtis dissimilarities (BCDs) with samples colored by study. In the MDS plot, the first two dimensions are shown, each with variation explained by the dimensions in parentheses. Statistical significance of sample grouping based on BCDs was determined using PERMANOVA, with p-value shown on bottom-right. **c)** Comparison of taxa (y-axis) log-2 fold changes (L2FCs, axis) between NPS and NPA samples. Each taxon is colored based on whether it was SDA (red) or not (grey), and the shape of the data point indicates whether the taxon passed sensitivity analysis (SA) (triangles) or not (circles). ANCOM-BC2 employs SA by evaluating whether results change if different pseudo-counts are used to replace zeros in the dataset (if so, then the taxon fails SA). Horizontal bars for each data point represent the 95% confidence interval of the L2FC measurement computed using its standard error. Positioning of the data points/bars to the right of 0 indicates an enrichment of taxa in NPS samples relative to NPA samples, while those to the left of 0 are depleted. Taxa are ordered by mean relative abundance (most abundant at the bottom of plot). **d)** Boxplots of two highly dissimilar taxa found in **c**, showing relative abundances (RAs) stratified by study (log10 axis transformation).

We first assessed overall diversity and composition. Alpha diversity was significantly higher in NPS samples compared to NPA (**Figure 4a**; Wilcoxon rank-sum test, W=87,737, *P*<0.0001), a pattern consistent across multiple rarefaction thresholds. Beta diversity analysis using Bray-Curtis dissimilarity showed significant differences in community structure between the two sampling methods (**Figure 4b**; PERMANOVA, *P*<0.001).

To identify differentially abundant taxa, we applied ANCOM-BC2 to the full dataset. Of the 632 taxa, 42 met prevalence thresholds for analysis, and 27 (64%) were significantly differentially abundant (SDA; *P*<0.05) (**Figure 4c**). Among these, 21 passed the sensitivity analysis. Notable differences included enrichment of *g_Moraxella* (L2FC=2.7) and *g_Haemophilus* (L2FC=2.3) in NPS samples, and *g_Staphylococcus* (L2FC=-4.3) in NPA samples. The species *s_Moraxella lincolnii* had the largest L2FC (5.7) but was excluded during sensitivity analysis, likely due to insufficient resolution in the V4 region. Further analysis revealed higher *g_Moraxella* and lower *g_Staphylococcus* relative abundance in NPS samples (**Figure 4d**). We also observed a strong co-exclusion pattern between *g_Moraxella* and *g_Staphylococcus*: in 70% of samples (494 of 706), only one was detected above 1% relative abundance (**Table 2**; Pearson’s Chi-squared test, *X*^*2*^=143.81, df=3, *P*<0.0001).

**Table 1.** Summary of NPS and NPA datasets analyzed.

| Study | NCBI BioProject | Sample type | Age range | Total samples | Age-matched, healthy, & filtered samples |
| --- | --- | --- | --- | --- | --- |
| **Odendall *et al* (19)** | PRJNA997934 | NPS | All | 2,799 | 317 |
| **Teo *et al* (20)** | PRJNA275918 | NPA | 2-12 m/o | 913 | 391 |

**Table 2.** Comparison of g_Moraxella and g_Staphylococcus detection between NPA and NPS samples, using a detection threshold of 1% relative abundance (RA).

| g_Moraxella RA > 1%? | g_Staphylococcus RA > 1%? | Number of NPA samples | Number of NPS samples |
| --- | --- | --- | --- |
| **No** | No | 90 | 52 |
| **No** | Yes | 133 | 12 |
| **Yes** | No | 121 | 228 |
| **Yes** | Yes | 45 | 25 |

Together, these findings demonstrate that the choice of sampling method significantly influences the diversity and taxonomic composition of the nasopharyngeal microbiome as recovered by sequencing.

## Discussion

Our analysis identified substantial gaps in reproducibility, usability, and integration, demonstrating how non-standardized reporting severely limits the scientific value and reusability of metagenomic datasets, undermining their potential to contribute to robust and replicable scientific discoveries.

### Reproducibility Challenges in Metagenomic Studies

Reproducibility, a cornerstone of scientific rigor, is severely hampered by inadequate reporting methods in nasopharyngeal metagenomic studies. In our study, only 78 out of 148 (52.7%) of the analyzed studies met the reproducibility criteria for methods sections, suggesting that many studies lack the necessary detail for other researchers to replicate their findings. This gap in reporting includes insufficient information on critical steps, such as DNA/RNA extraction found to be missing in 22 (14.9%) studies, sequencing protocols missing in 12 (8.1%) studies, source designation missing in 26 (17.5%) studies, and sample collection missing in 106 (71.6%) studies, which are essential to ensure that results can be independently verified. The lack of detail in these areas creates uncertainty around reported findings, limits trust in conclusions drawn from the data, and makes it challenging for researchers to build on prior studies. Improving reproducibility through standardized and comprehensive reporting would enable more reliable comparisons across studies, advancing our understanding of microbial ecology in the nasopharynx.

### Usability Constraints Due to Incomplete Metadata

Inconsistent reporting also limits the usability of data for new research contexts, with metadata availability emerging as a significant challenge. Of the 78 studies with reproducible methods sections, only 33 (42.3%) had usable metadata in public datasets. Metadata—information that describes the context, conditions, and parameters of an experimental dataset—is essential for researchers looking to reanalyze or apply data beyond the scope of the original study. In the absence of comprehensive and accessible metadata, the usability of data is compromised, as researchers lack the contextual information necessary to conduct accurate analyses or to interpret results meaningfully. To maximize the potential of metagenomic data, we recommend that journals and repositories enforce standardized metadata submission that includes, at minimum, clear subject demographics, health status, sampling protocols, and experimental conditions.

Furthermore, our findings revealed that 3.1% of nasopharyngeal metagenomic studies had mismatched laboratory methods, where the sequencing methods reported did not correspond with the actual data formats in public repositories. This misalignment further complicates usability, as it reduces the reliability of the data and raises fundamental questions about the accuracy and validity of study conclusions. Standardized protocols for both reporting methods and metadata alignment are critical for ensuring that publicly available datasets are usable and accurately reflect the experimental conditions under which they were generated.

### NPS Versus NPA Sampling for the Nasopharyngeal Source

Comparative analysis of nasopharyngeal swab (NPS) and aspirate (NPA) samples revealed significant differences in microbial diversity and community composition, with profound implications for reproducibility in nasopharyngeal microbiome research. We identified *g_Moraxella, g_Haemophilus*, and *g_Staphylococcus* as key taxa differentially abundant between the two sampling methods, suggesting that sampling technique plays a critical role in shaping observed microbiome profiles.

One of the most striking findings was the markedly higher abundance of *g_Staphylococcus* in NPA samples (L2FC = –4.3), indicating more than a 16-fold increase relative to NPS samples. This substantial enrichment likely reflects the increased capture of oral-associated microbes during aspiration-based collection. Unlike swabbing, which typically targets the posterior nasopharynx with minimal disturbance to surrounding areas, aspirates may collect a broader range of microorganisms from the upper respiratory tract, including the oropharyngeal space.

*g_Moraxella* and *g_Haemophilus* were significantly enriched in NPS samples, and both taxa are considered common and clinically relevant colonizers of the nasopharynx. Their higher relative abundance in swab-collected samples may suggest that NPS more effectively targets the proper nasopharyngeal niche, while aspirates may dilute this signal with oral contaminants. Similarly, although less pronounced, *g_Rothia* also showed a trend toward differential abundance, further supporting the influence of sampling method on community structure.

These findings underscore the critical importance of standardized sampling protocols in nasopharyngeal microbiome research. The apparent compositional disparities observed between NPA and NPS methods emphasize the need for meticulous documentation of collection procedures. Without a consistent methodology, cross-study comparisons may be confounded by sampling artifacts, potentially leading to misleading conclusions in research and clinical contexts. As microbiome-based diagnostics and interventions gain traction, avoiding methodological bias through standardized collection protocols and transparent reporting is essential to ensure accuracy and reproducibility.

### Implications for Data Integration Across Studies

Integration of data from multiple studies is essential for large-scale analyses that drive a comprehensive understanding of metagenomics, but the inconsistent reporting practices we identified severely hinder this process. Data integration requires that methodologies, metadata, and laboratory procedures align across studies for reliable aggregation and comparative analyses. The discrepancies in reproducibility, metadata usability, and method compatibility identified in this study highlight the challenges in combining datasets for more expansive research opportunities. Misaligned or inadequately reported methods introduce variability and may lead to spurious results when datasets are integrated. For example, combining NPS and NPA datasets without accounting for their inherent microbial profiling differences could yield artificial clusters in meta-analyses that reflect methodology rather than biology. Adopting standardized reporting protocols that detail sample collection, sequencing, and data processing methods is essential to support meaningful data integration necessary for larger initiatives, such as population-scale microbiome-disease association studies and respiratory pathogen surveillance.

### Recommendations and Future Directions

Our findings underscore the pressing need for standardized reporting guidelines in nasopharyngeal metagenomics, which could potentially have broader applications in other metagenomic fields. Consistent criteria for reporting methodological details, metadata, and data alignment practices would benefit reproducibility, usability, and integration across studies.

To address the reproducibility challenges identified in our review, we propose a comprehensive framework for standardized reporting in metagenomic research. Metagenomic sequencing is an invaluable tool for studying the complex microbial communities that inhabit diverse environments, from human microbiomes to soil ecosystems. However, ensuring the reproducibility and replicability of metagenomic research requires detailed and transparent reporting of sequencing data. As a best practice, provide comprehensive metadata that covers the “how,” “why,” and “where” of data generation, including technical and procedural details, ranging from sample collection through bioinformatic analysis protocols. Based on our systematic review findings, we have identified several critical reporting elements that should be considered essential in nasopharyngeal research publications. These elements generally fall into five key areas: 1) Sequencing platform and technology, 2) Sample processing and DNA extraction, 3) Library preparation and sequencing depth, 4) Bioinformatics and quality control, and 5) Contextual metadata.

The sequencing platform chosen for metagenomic studies significantly impacts the quality, depth, and data type obtained. Different sequencing platforms come with their advantages and limitations, so understanding these details is essential for reproducibility and comparing results across studies. Researchers should identify the sequencing platform used, such as Illumina, Oxford Nanopore, PacBio, or Roche 454, and specify the exact model (e.g., Illumina NovaSeq, MiSeq) and the manufacturer. Each platform has inherent biases, read lengths, and error profiles that can influence the sequencing outcomes. For example, Illumina platforms generate short, high-accuracy reads, while Oxford Nanopore offers longer reads better suited for genome assembly but with a higher error rate. The sequencing technology should also be specified, whether next-generation sequencing (NGS) or third-generation sequencing (TGS). As a best practice, mention whether shotgun sequencing (entire genome sequencing) or targeted sequencing (e.g., 16S rRNA gene sequencing for microbial identification) was employed, as these methods produce different data types and require distinct analysis approaches. The sequencing chemistry or library preparation protocols should also be detailed, including whether single-end or paired-end reads were generated. These factors are crucial to understanding the data and ensuring results can be replicated across different studies.

Describing the DNA extraction protocol used, including the specific kit and any modifications, is crucial. DNA extraction methods can introduce biases in microbial recovery, especially in complex microbial communities. Some methods may preferentially extract DNA from specific taxa or fail to recover DNA from particular organisms. Additionally, researchers should specify the library preparation kit employed, as different kits can affect the amplification or fragmentation of DNA. The library preparation process plays a pivotal role in shaping the quality and structure of the sequencing data. Proper documentation of the library preparation protocols helps ensure that other researchers can replicate the procedure and assess its impact on data quality and microbial profiling. For example, high-throughput sequencing kits may perform differently than those optimized for low-biomass samples. Furthermore, researchers should explain how DNA or library concentration was measured and normalized before sequencing, as variations in library concentration can lead to sequencing depth imbalances across samples. Proper reporting of these protocols is essential, as biases in library preparation can significantly influence microbial diversity estimates and taxonomic classifications.

Sequencing depth, or the number of reads generated per sample, directly affects the resolution and accuracy of microbiome characterization. Along with sequencing depth, read quality is a key factor in ensuring the reliability of the sequencing data. Researchers should clearly report the number of samples that failed to meet predefined quality thresholds and explicitly describe these criteria in the methods section. Furthermore, samples that do not meet minimum standards should not be uploaded to public repositories unless accompanied by comprehensive metadata that clearly indicates their quality status and limitations. Higher sequencing depth increases sensitivity, allowing for the detection of rare microbial taxa, while shallow sequencing may miss low-abundance species, limiting the ability to describe microbial diversity accurately. The read length should also be specified, as it can differ depending on the platform (e.g., 150 base-pair paired-end reads on Illumina versus longer reads from PacBio or Oxford Nanopore). Longer reads provide more comprehensive genomic information, whereas shorter reads may be more suitable for targeted sequencing, like 16S rRNA gene sequencing, but may result in less accurate taxonomic assignments in shotgun sequencing. It is also important to detail the quality control measures applied to ensure the integrity of the sequencing data. These measures may include filtering out low-quality reads, adapter sequences, and host contamination, such as human DNA, that could interfere with microbiome analysis. Researchers should specify the software and parameters used to filter out low-quality sequences, such as read length thresholds, Phred score cutoffs, or quality trimming procedures. Insufficient read depth or poor-quality data can compromise the accuracy of taxonomic and functional analyses, so transparency in these procedures allows other researchers to assess whether the data meets the necessary standards for reproducibility.

The bioinformatics pipeline used to process sequencing data is crucial in shaping the results, especially in microbial identification, taxonomic classification, and functional annotation. Clear documentation of the data processing steps is essential for reproducibility. Researchers should report all preprocessing steps, including trimming, quality filtering, removal of adapter sequences, and filtering of host genome sequences. These steps ensure the data is consistently prepared, minimizing potential biases during downstream analysis. Details about the taxonomic classification software or algorithm used should be provided. Different databases and classification methods may yield different results, especially when distinguishing between closely related microbial species or strains. If functional analysis was conducted, researchers should explain the software and databases used for functional annotation, as well as any statistical methods employed for functional annotation and enrichment analysis. Researchers should also specify any normalization techniques applied, such as rarefaction or total-sum scaling, and the statistical methods used for group comparisons, such as PERMANOVA, ANOVA, or differential abundance analysis. Detailed reporting of these bioinformatics steps ensures that any biases introduced during sequencing or preprocessing do not distort the results and allows other researchers to replicate the analysis.

Beyond the technical aspects of sequencing, it is crucial to provide contextual metadata that helps to interpret the results accurately. This includes describing the geographical location, environment, and ecological context of the sample, such as whether it comes from the human gut, oral cavity, or ocean water, as these factors can heavily influence microbial composition. Additionally, the experimental design should be documented in detail, including the number of replicates, control groups, and any treatments or interventions, such as antibiotic exposure or dietary changes. For longitudinal studies, it is important to report the timepoints of sample collection to account for temporal changes in microbial communities. Providing this contextual information ensures that the sequencing data can be properly interpreted and accurately compared with other studies, helping researchers understand the factors that could influence microbial composition.

In conclusion, while metagenomic sequencing offers significant insights into microbial ecosystems, the ability to replicate and compare findings depends on clear and thorough reporting of sequencing data. By documenting critical details, including sequencing platforms, library preparation protocols, sequencing depth, data processing methods, and experimental design, researchers ensure that their findings are reproducible and comparable. This level of transparency allows for meaningful cross-study comparisons and the advancement of our understanding of microbial communities across various environments.

Initiatives such as the Minimum Information About a Metagenomic Experiment (MIMS), Genomic Standards Consortium (GSC), and the National Institute of Standards and Technology (NIST) provide a starting point for developing robust guidelines, and adherence to such standards could be encouraged by journals and funding agencies. By requiring complete and standardized reporting, the metagenomic community can improve the transparency and utility of published studies. Standardization acts as a firm foundation for continuing progress and expansion within the field.

### Limitations and Scope for Future Research

This systematic review focused specifically on nasopharyngeal metagenomic datasets and their suitability for integration into a unified bioinformatics resource. While the findings highlight critical gaps in data quality, reproducibility, and reporting, they may not be generalizable to metagenomic studies involving other anatomical sites or sample types. Moreover, a key limitation of this review is the lack of paired sample collection; nasopharyngeal aspirates and swabs were not collected from the same subjects across studies, introducing potential confounding variables such as host age, comorbidities, environmental exposures, and geographic location that may influence observed microbial differences.

Future research should explore whether similar inconsistencies in metadata reporting and methodological alignment exist in broader metagenomic research contexts. Additionally, further investigation is needed to elucidate the underlying causes of methodological mismatches and insufficient metadata, which may guide the development of targeted, field-wide standards to enhance the utility of publicly available datasets. Developing automated tools for metadata quality assessment and standardization would greatly benefit the microbiome research community.

## Conclusion

In conclusion, our systematic review reveals significant challenges in the reproducibility, usability, and integration of nasopharyngeal metagenomic datasets due to insufficient and inconsistent reporting practices. With only 34.3% of studies meeting reproducibility criteria, 42.36% of those studies providing usable metadata, and 3.1% showing mismatched methods, it is evident that standardized reporting guidelines are urgently needed to advance the reliability and utility of metagenomic research. Strikingly, only 4.6% (33 accepted of the 710 screened) of publicly available datasets were ultimately suitable for advancing the definition of a healthy nasopharyngeal microbiome, highlighting the substantial scientific opportunity cost of poor reporting practices. Our study also highlights how the nasopharyngeal sampling method significantly impacts microbiome composition. Researchers should exercise caution when interpreting results across studies using different collection techniques and prioritize transparent reporting of methodological details. By implementing the standardized reporting practices we outlined, the field can build a stronger foundation for reproducible, usable, and integrative research. Ultimately, improved standardization will enhance the scientific value and clinical impact of metagenomic studies, accelerating discoveries that refine our understanding of the nasopharyngeal microbiome.

## Data Availability

Raw 16S V4 rRNA gene sequencing data were obtained from NCBI BioProject PRJNA997934 (nasopharyngeal swabs, NPS) and 16S V3V4 rRNA gene sequencing data from PRJNA275918 (nasopharyngeal aspirates, NPA).

https://www.ncbi.nlm.nih.gov/sra

## Author Contributions

**Monica Bustos**: Conceptualization; data curation; formal analysis; supervision; project administration; resources; software; validation; visualization; writing - original draft; writing - review & editing. **Kuncheng Song:** Conceptualization; Data curation; formal analysis; visualization, methodology; resources; software; writing - review & editing. **Hayden N Brochu:** Data curation; formal analysis; methodology; visualization; resources; software; writing - review & editing. **Qimin Zhang:** Data curation; resources; **Lakshmanan K Iyer**: writing - review & editing. **Crystal R Icenhour**: Conceptualization; Supervision; project administration; writing - review & editing.

## Acknowledgements

All authors are current or former employees of Labcorp, a provider of clinical diagnostic services. We would like to thank the U.S. Centers for Disease Control and Prevention (CDC) for their guidance and expertise throughout the REaSoN project.

